# Tenzingplasty for Cerebral Vasospasm Following Subarachnoid Hemorrhage and Meningitis: Multicenter Initial Experience

**DOI:** 10.1101/2025.05.12.25327468

**Authors:** MD Alexander, N Mehta, F Behzadi, WT Kim, J Kim, N Telischak, RS Khangura, B Varjavand, TA Chaudhry, F Settecase, JD English

## Abstract

Cerebral vasospasm is a morbid complication of intracranial pathologies. Heterogeneity exists in treatment regimens, but management usually involves intensive medical therapy that may require augmentation with endovascular therapy, most commonly intra-arterial CCB infusion; angioplasty is historically reserved for refractory cases with a high risk for permanent neurological deficits. Given the high risk of iatrogenic complications with balloon angioplasty, alternative mechanical therapies have been explored. This series examines Dotter technique mechanical dilatation performed with Tenzing devices (Tenzingplasty).

**Methods:** IRB-approved retrospective analysis of prospectively maintained procedural databases and medical records was performed for patients undergoing Tenzingplasty at five high-volume cerebrovascular centers. Demographics, clinical features, and treatment details were recorded. Per-segment analysis was performed, including any treated arterial segment with narrowing ≥50%. The primary outcomes were improvement of treated vessel narrowing and improvement to less than 50% of baseline diameter. The primary safety outcome was any procedural complication. Secondary outcomes included absolute improvement in narrowing and need for repeat mechanical endovascular therapy to be performed on a previously targeted segment. Statistical analysis was performed with the Exact test for non-continuous variables and T-tests for continuous variables. Mixed effects linear and logistic regression analysis was then performed as appropriate for multivariable analysis.

**Results:** Fourteen patients were treated with Tenzingplasty for vasospasm; 12 (85.7%) had aSAH, and 2 (14.3%) had meningitis. Tenzing devices were passed through 82 arterial segments with narrowing ≥50%. All treated segments had improved narrowing, with 78 (95.1%) having residual narrowing <50%. No procedural complications occurred. Mean (±SD) vessel narrowing improved from 80±13% to 34±14%, with absolute improvement of narrowing 46±17%. Three (3.7%) segments required repeat mechanical endovascular therapy. In univariable and multivariable analysis, no demographic, clinical, or treatment variables were associated with any

**Conclusion:** Tenzingplasty performed for cerebral vasospasm was safe and effective, with vessel narrowing improving in all treated segments. No procedural complications occurred in any treated segment. Low need for repeat mechanical endovascular therapy suggests a durable effect.

Further investigation is warranted.

## Introduction

Cerebral vasospasm (CV) is a well-known complication following aneurysmal subarachnoid hemorrhage (aSAH) and intracranial infections that can be difficult to manage and may lead to potentially devastating outcomes despite available aggressive medical and endovascular treatment. The pathophysiology of CV is complex and incompletely characterized, known to involve activation of both inflammatory and ischemic cascades that can result in delayed cerebral ischemia (DCI).[1,2] Numerous treatment approaches have been proposed with variable efficacy, and most treatment modalities have not been adequately studied with rigorous prospective investigations.[1] As such, treatment algorithms often vary dramatically across geographical regions, medical centers, and treating providers.

Medical management of CV traditionally includes hemodynamic measures like intravenous fluids for volume expansion, vasopressors to augment cerebral perfusion with higher blood pressure, and calcium channel blockers (CCB) to deactivate voltage-gated calcium channels in smooth muscle cells of arterial walls, either orally (nimodipine) and/or intra-arterially (verapamil).[3,4] When these measures fail, more aggressive endovascular intervention can be pursued with mechanical dilatation of arteries. This has historically been performed with balloon angioplasty.[3,5] While typically efficacious for improving arterial caliber, balloon angioplasty can be technically challenging and carries a high risk for dissection, thromboembolic complications, or vessel rupture. Vessel rupture in particular is highly morbid and can be fatal; this feared complication often prevents operators from pursuing angioplasty until CV is clearly refractory to IA CCB infusion and likely to result in DCI. In recent years, operators have explored alternative mechanical strategies for CV relief, such as stent-retrievers, including the NeVa device (Vesalio, Nashville, TN), or stent-retriever like devices such as the Comaneci device (Rapid Medical, Yokneam, Israel).[6–8]

Charles Dotter first reported catheter-mediated mechanical dilatation in a peripheral arterial atherosclerotic stenosis in 1964.[9] This has come to be referred to as the Dotter technique, or Dotterization. It has not previously been reported for the intracranial arteries. The Tenzing device (Route 92 Medical, San Mateo, CA) is a delivery catheter designed for delivering aspiration catheters for mechanical thrombectomy; its design includes a soft, atraumatic, flexible, lubricious tip with a tapered design for ledge reduction.[10–14] The Tenzing tip may be suitable for Dotter angioplasty of intracranial atherosclerotic disease, with early reports of favorable outcomes using the Tenzing device to recanalize acutely occluded atherosclerotic lesions among initial case series.[10–15] The design features of the Tenzing device may also render it suitable for Dotter angioplasty in the setting of severe CV. The Tenzing device currently comes in 3 sizes, with a maximal outer diameter (OD) of 1.2 mm in the Tenzing 5, 1.6 mm in the Tenzing 7, or 2.1 mm in the Tenzing 8 (Figure 1). The Tenzing OD approximates the diameter of the proximal intracranial arteries and is comparable to angioplasty balloon inflation sizes used for refractory CV.[16–18] In this study, we evaluate the feasibility, efficacy, and safety of novel use of the Tenzing device for performance of the Dotter angioplasty — Tenzingplasty—for CV.

**Figure 1.**
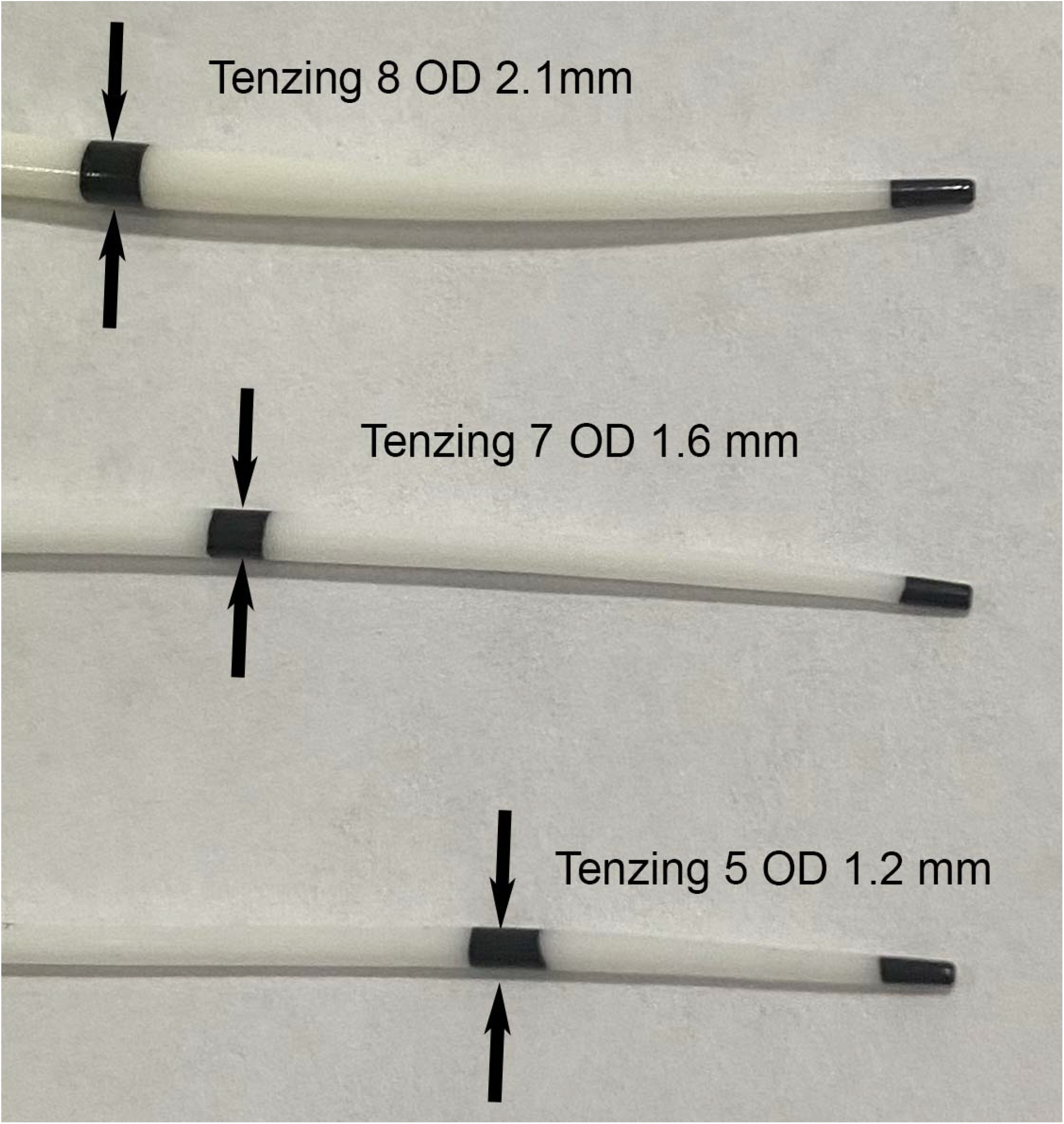
Photograph of Tenzing 8, Tenzing 7, and Tenzing 5 device tips with maximal OD at proximal markers indicated by arrows.

## Methods

### Study Design and Patient Population

After IRB-approval, a search of prospectively maintained institutional databases was performed for patients undergoing Tenzingplasty for CV at five high-volume neurointerventional centers was performed. Inclusion criteria for Tenzingplasty treatment included CV necessitating mechanical treatment after failure of noninvasive medical therapy and intra-arterial infusion of CCBs. Data were retrospectively collected, including demographic characteristics, etiology of CV (aSAH vs. meningitis), Hunt & Hess and modified Fischer scores for aSAH patients, and procedural details. Per-segment analysis was performed, including any arterial segments treated with Tenzingplasty for ≥50% narrowing compared to baseline segment diameter.

### Tenzingplasty Procedural Details

Tenzingplasty was performed by advancing the Tenzing device over a 0.014” microwire under fluoroscopic guidance. Intravenous heparin was administered in most cases, seeking to double baseline activated clotting time. The Tenzing device size was chosen at the discretion of the operator, selecting the device with OD that would best approximate but not exceed the normal segment diameter on premorbid angiography. Choice of guide catheter and microwire were chosen at the discretion of the operator. Under direct fluoroscopic visualization and roadmap guidance, the device was slowly advanced so the proximal radiopaque marker, at which point the tapered tip reaches maximum OD, was passed to the point of interest. (Figures 2 and 3) For instance, a Tenzing 5 would be advanced into M2 and M3 branches for CV in medium vessels, and a Tenzing 7 would then be advanced with the proximal marker into the proximal M2 segment for better result in the M1 segment. (Figures 2 and 3) The device was advanced to its target in one smooth, continuous movement. Dwell time and administration of CCBs were left to operator discretion. Dwell time was typically 1-60 seconds, and intra-arterial infusion could be performed before the pass, after the pass, or during dwell time by injecting through the lumen of the Tenzing device. After the Tenzingplasty pass, angiography was immediately performed by injecting the guide catheter from its upstream position.

**Figure 2.**
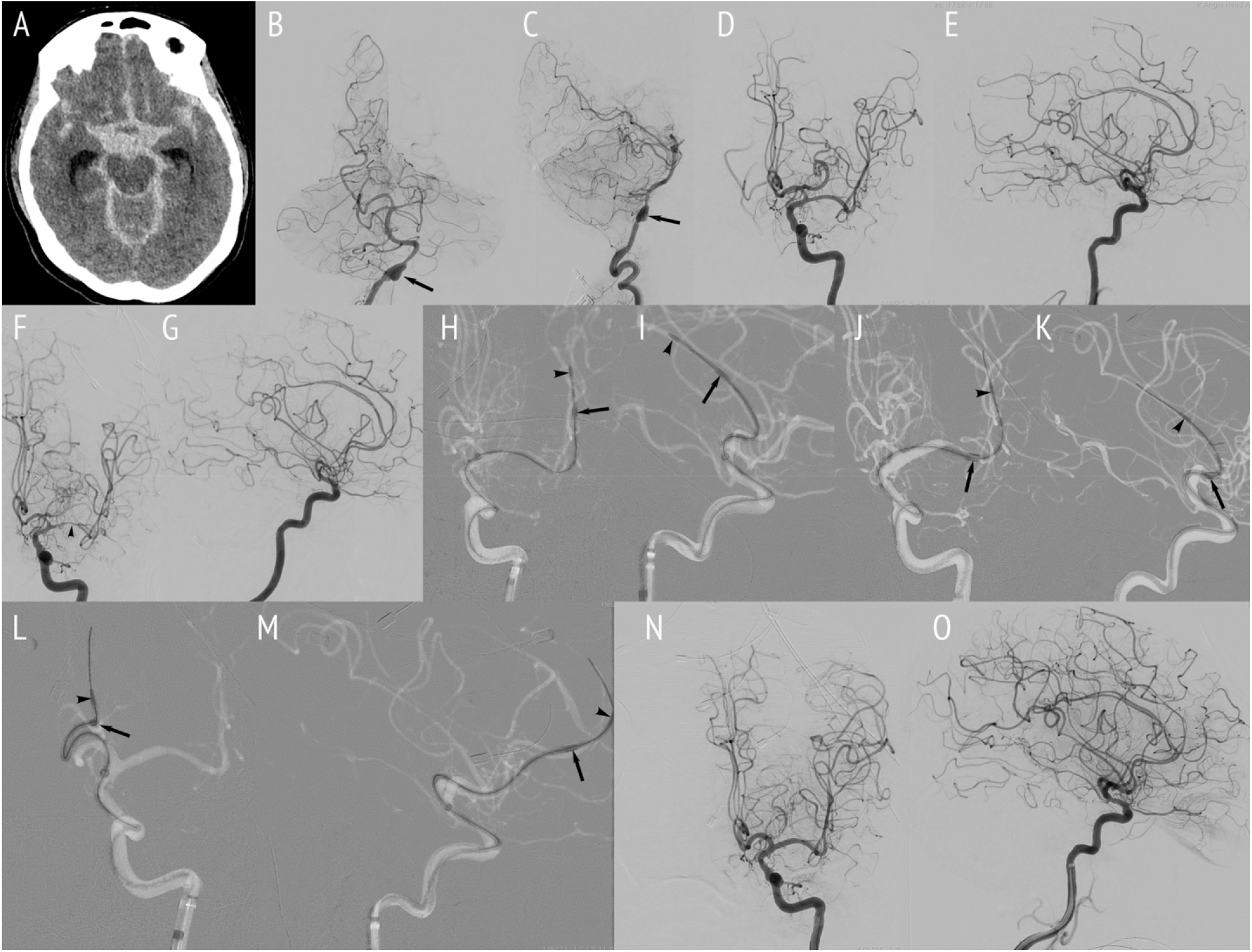
A female patient in her early 60s presented with Hunt & Hess 5 modified Fisher 4 aSAH (A) from a ruptured dissecting aneurysm (arrows) of the right vertebral artery, seen in Towne’s (B) and lateral (C) projections. Initial Towne’s (D) and lateral (E) projections during injection of the left ICA show no vasospasm on bleed day 1 when the rupture site was treated with a flow-diverting stent. Frontal (F) and lateral (G) projections of the left ICA on bleed day 5 show severe multifocal vasospasm, most pronounced in the left M1 segment (arrowhead). Tenzingplasty was performed over a Synchro microwire in multiple branches, with arrowheads indicating distal markers and arrows indicating proximal markers (H-M). A Tenzing 5 device was used for treatment in left M3 branches (H, I). A Tenzing 7 device was used to treat left M1-2 segments (J, K) and A1-2 segments (L, M). Post-treatment injection of the left ICA (N, O) demonstrates improved caliber in treated segments.

**Figure 3.**
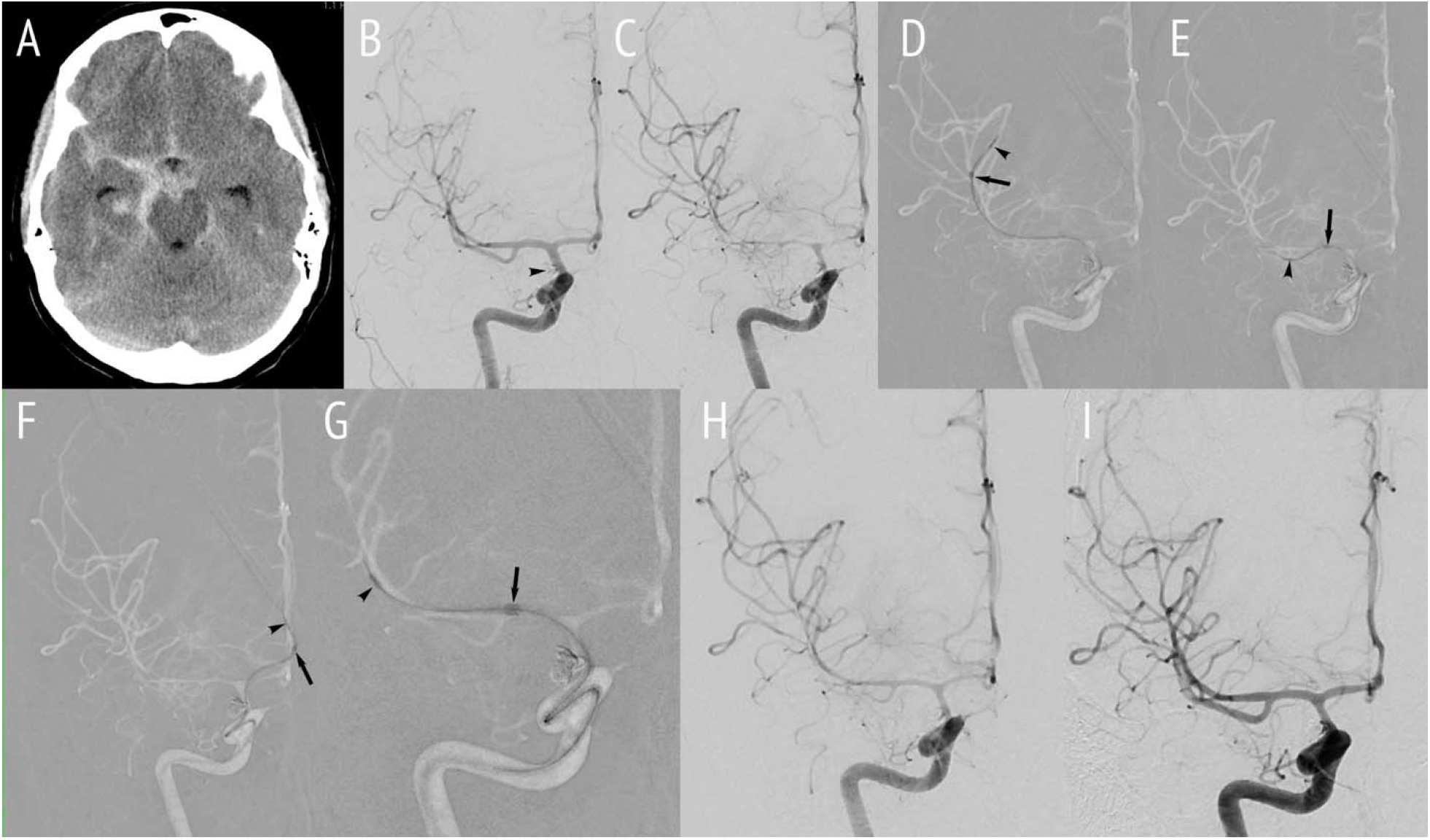
A female patient in her mid-40s presented with Hunt & Hess 2 modified Fisher 3 aSAH (A) from a ruptured right posterior communicating artery aneurysm, which was coiled (arrowheads) on bleed day 1. Neurological deficits and elevated TCD velocities prompted angiography (C) on bleed day 5, which demonstrated multifocal vasospasm in the right ICA, MCA, and ACA. Tenzingplasty was performed (D-G), with arrowheads indicating distal tips and arrows indicating proximal markers of the devices, which were advanced over a Synchro wire. A Tenzing 5 device was used for treatment in the posterior (D) and anterior (E) divisions of the right MCA and the A2 segment of the right ACA (F). A Tenzing 7 device was used for treatment of the right M1 segment (G). Post-treatment angiography (H) demonstrates improved vessel caliber. Repeat angiography on bleed day 8 demonstrated durable response, with no mechanical treatment required for any of the previously treated segments.

### Efficacy and Safety Endpoints

The degree of pre vs post Tenzingplasty stenosis was calculated based on the region of worst narrowing of the arterial segment on the pre-treatment high magnification angiographic run at the time of CV, compared to the normal caliber of the affected segment from the angiography performed earlier during the patient’s admission prior to the development of CV. If premorbid imaging was not available, which occurred in patients with meningitis who presented after CV had begun, postmorbid vascular imaging after resolution of CV was evaluated to determine baseline diameters. Primary outcomes were any improvement in narrowing of the treated segment, either in absolute measurement or according to the ordinal scale utilized in the COMMAND study, and residual narrowing <50% immediately after treatment, mirroring VITAL methodology.[6,7] When more than one Tenzing size was used for treatment, post-treatment measurements following Tenzingplasty with the largest device were used. The primary safety endpoint was any procedural complication. Secondary outcomes included total reduction in narrowing following Tenzingplasty and need for repeat endovascular mechanical treatment of CV.

### Statistical Analysis

Statistical analysis was performed in STATA version 17 (StataCorp, College Station, TX). Summary statistics were performed for demographic variables and clinical features. Univariable analysis was performed with t-tests for continuous variables and the Fisher-Freeman-Halton exact test for non-continuous variables. Mixed effects linear regression analysis was then performed for multivariable analysis, excluding variables with p>0.20 in univariable tests to account for possible residual confounding.

## Results

Fourteen patients underwent Tenzingplasty: 12 (85.7%) with aSAH and 2 (14.3%) with meningitis. Table 1 summarizes demographic and clinical presentation variables; Table 2 summarizes such clinical variables for aSAH patients. A total of 82 arterial segments with ≥50% narrowing were treated and included in the analysis. A total of 92 Tenzing passes were made in these segments, with no dissection, perforation, or occlusion observed. Twelve (23.9%) passes were made with Tenzing 5, 42 (45.7%) with Tenzing 7, and 23 (25.0%) with Tenzing 8. No immediate re-treatment was needed in any cases, although some repeat passes were made after the operator decided to repeat Tenzingplasty to treat further distally than during the initial pass. Table 3 summarizes pre- and post-treatment diameters of arterial segments with post-treatment measurements delineated by device.

**Table 1:** Demographic and Clinical Features.

|  |  |
| --- | --- |
| Biological Sex |  |
| Female | 7 (50.0%) |
| Male | 7 (50.0%) |
| Age (Years ±SD) | 50.8 ± 10.4 |
| Cause of Vasospasm |  |
| aSAH | 12 (85.7%) |
| Meningitis | 2 (14.3%) |
| Comorbidities |  |
| Hypertension | 6 (42.9%) |
| Hyperlipidemia | 5 (35.7%) |
| Coronary Artery Disease | 5 (35.7%) |
| Tobacco Use | 5 (35.7%) |
| Previous Cocaine Use | 2 (14.3%) |
| Previous Amphetamine Use | 3 (21.4%) |

**Table 2:**
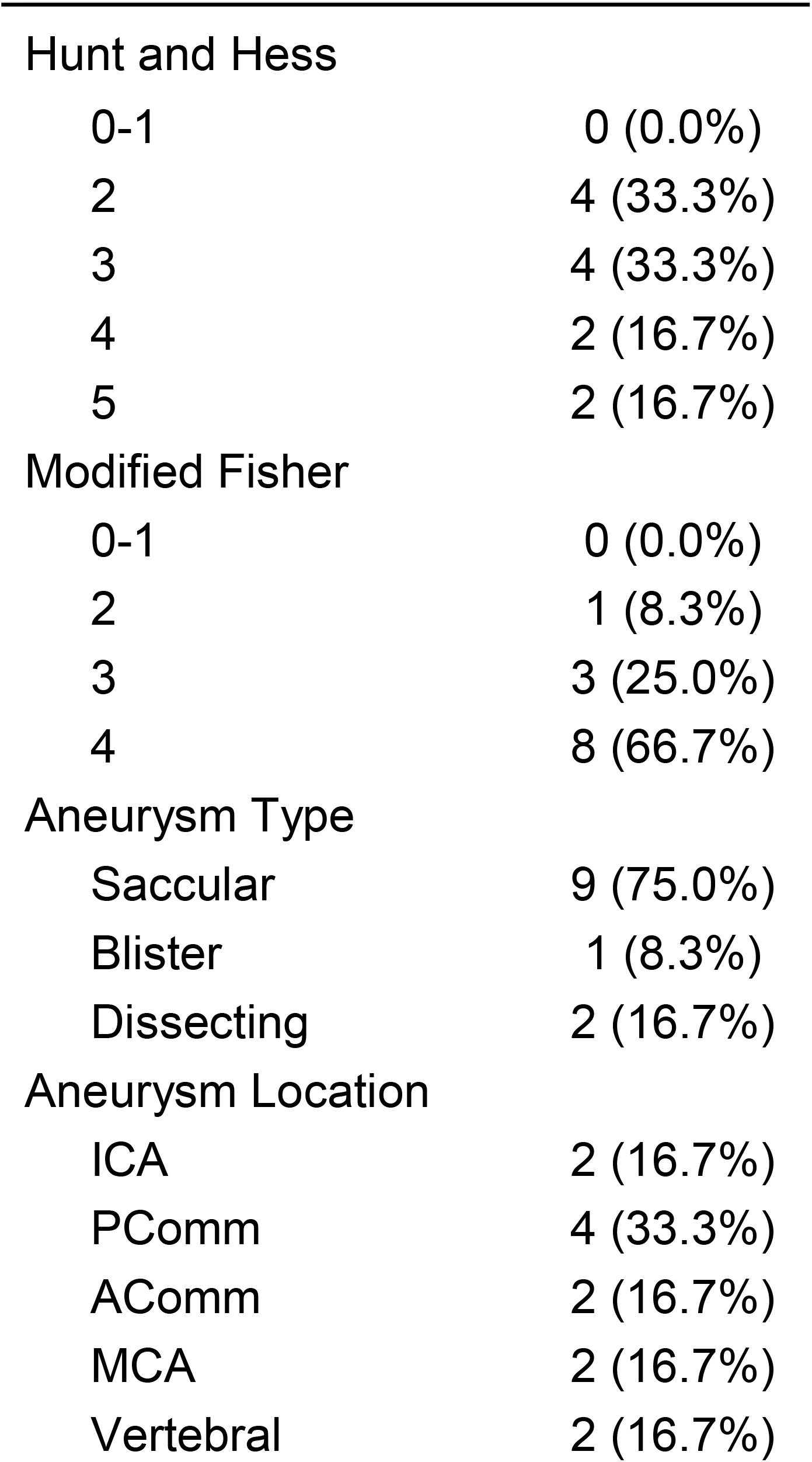
Aneurysm Presentation Details (n=12)

|  |  |
| --- | --- |
| 0-1 | 0 (0.0%) |
| 2 | 4 (33.3%) |
| 3 | 4 (33.3%) |
| 4 | 2 (16.7%) |
| 5 | 2 (16.7%) |
| Modified Fisher |  |
| 0-1 | 0 (0.0%) |
| 2 | 1 (8.3%) |
| 3 | 3 (25.0%) |
| 4 | 8 (66.7%) |
| Aneurysm Type |  |
| Saccular | 9 (75.0%) |
| Blister | 1 (8.3%) |
| Dissecting | 2 (16.7%) |
| Aneurysm Location |  |
| ICA | 2 (16.7%) |
| PComm | 4 (33.3%) |
| AComm | 2 (16.7%) |
| MCA | 2 (16.7%) |
| Vertebral | 2 (16.7%) |

**Table 3:** Pre- and Post-Treatment Diameter and Narrowing by Segment and Device (n=12)

| Segment | n | Baseline<br>Minimal<br>Diameter | Vasospasm<br>Minimal<br>Diameter | Vasospasm<br>Narrowing | Post-T5<br>Minimal<br>Diameter | Post-T5<br>Narrowing | Post-T7<br>Minimal<br>Diameter | Post-T7<br>Narrowing | Post-T8<br>Minimal<br>Diameter | Post-T8<br>Narrowing |
| --- | --- | --- | --- | --- | --- | --- | --- | --- | --- | --- |
| ICA | 8 | 3.1 ± 0.9 | 1.0 ± 0.3 | 80.4% ± 17.1 | 1.7 ± 0.0 | 50% ± 0.0 | 1.8 ± 0.3 | 41.2% ± 4.4 | 2.4 ± 0.2 | 0.0% ± 0.0 |
| A1 | 6 | 1.7 ± 0.4 | 0.3 ± 0.1 | 83.0% ± 8.2 | 0.9 ± 0.3 | 40.0% ± 9.1 | 1.4 ± 0.3 | 16% ± 13.3 | - | - |
| A2 | 5 | 1.5 ± 0.5 | 0.3 ± 0.2 | 80.5% ± 8.7 | 1.1 ± 0.1 | 24.6% ± 28.3 | 1.2 ± 0.2 | 17.5% ± 24.7 | - | - |
| A3 | 5 | 1.4 ± 0.5 | 0.3 ± 0.1 | 74.5% ± 18.2 | 1.0 ± 0.0 | 0.0% ± 0.0 | 1.3 ± 0.3 | 15.8% ± 13.9 | - | - |
| M1 | 18 | 2.1 ± 0.6 | 0.6 ± 0.3 | 77.1% ± 14.7 | 1.2 ± 0.4 | 41.7% ± 21.9 | 1.4 ± 0.2 | 32.1% ± 13.4 | 2.0 ± 0.4 | 24.2% ± 12.1 |
| M2 | 22 | 1.7 ± 0.7 | 0.5 ± 0.2 | 75.3% ± 14.2 | 1.0 ± 0.2 | 31.9% ± 19.6 | 1.3 ± 0.3 | 17.2% ± 12.8 | 1.8 ± 0.2 | 32.1% ± 20.1 |
| M3 | 15 | 1.3 ± 0.3 | 0.3 ± 0.1 | 77.6% ± 6.3 | 1.0 ± 0.1 | 23.7% ± 12.0 | 1.8 ± 0.0 | 0.0% ± 0.0 | - | - |
| P1 | 3 | 1.4 ± 0.4 | 1.0 ± 0.4 | 37.3% ± 10.4 | - | - | - | - | 1.5 ± 0.1 | 0.0% ± 0.0 |

All treated segments demonstrated angiographic improvement after Tenzingplasty, both in absolute measurement and in ordinal scale classification, with 78 (95.1%) showing <50% residual narrowing. Following Tenzingplasty, mean (+/-SD) vessel stenosis improved from 80±12.7% to 34±14%, with absolute improvement of narrowing 46±17%. Three (3.7%) segments required repeat mechanical endovascular therapy, which was performed with repeat Tenzingplasty for all three segments. No patient-specific or treatment variables were predictive of the primary effectiveness or safety outcomes in univariable or multivariable analysis.

In analysis of secondary outcomes, reduction in narrowing following treatment was associated with more distal arterial segments (p<0.001), Hunt and Hess score (p<0.001), modified Fisher score (p<0.001), symptoms referable to the segment undergoing treatment (p<0.001), patient age (p<0.001), hyperlipidemia (p=0.0083), coronary artery disease (p=0.0083), and history of tobacco (p=0.0398), cocaine (p<0.001), or amphetamine use (p=0.003). No variables were associated with the need for repeat endovascular mechanical treatment or durability on repeat imaging. None of these associations persisted in multivariable analysis.

## Discussion

CV is a highly morbid complication of aSAH or meningitis that can lead to DCI. In refractory cases, mechanical expansion of affected segments can be performed with angioplasty.[3,5] However, this procedure carries high treatment risk and is thus reserved for patients at high risk for DCI.[3,5] Given the inherent risk of angioplasty, alternative EVT approaches to vasospasm have recently been described using devices other than balloons, such as currently available stent-retrievers, the Comaneci device, and the NeVA device.[6–8] The current study demonstrates the feasibility, efficacy, and safety of Tenzingplasty as an alternative mechanical endovascular treatment for refractory CV. Tenzingplasty of CV resulted in significant improvement in arterial narrowing due to CV, with no complications. Few of the patients required repeat EVT, indicating a high degree of durability of Tenzingplasty. This is the first report of this novel technique for CV treatment.

Compared to traditional balloon angioplasty or more recently reported treatment with the Comaneci or NeVa devices, the Tenzing device allows for a more controlled, lower-profile, and potentially safer means of mechanical dilatation. Tenzingplasty may offer several advantages over traditional balloon angioplasty. Tenzingplasty treatment effect can be achieved on all segments through which a Tenzing device is passed, whereas balloon PTA, Comaneci, or NeVa treatment effect can only be achieved at sites where the device is inflated or deployed. Tenzingplasty can achieve its result with a single pass through affected vessels, whereas angioplasty carries a risk for complications with each inflation of a balloon, which must often be performed on multiple segments to achieve adequate results due to shorter balloon lengths. The same is true for stent-retrievers or the Comaneci device, which must be inflated at multiple sites or dragged while open to treat adjacent segments. The fixed sizes of Tenzing devices obviates variability in balloon diameter during dilation, need for choosing the degree of expansion with the Comaneci device, or excessive stress that may be applied to a vessel by the self-expanding design of a stent-retriever. Furthermore, the trackability of the Tenzing device allows for navigation to segments that would be more difficult and at higher risk of complications with balloon microcatheters or other devices, such as the A1 segment and more distal anterior and middle cerebral artery branches.

Compared to the COMMAND study, which evaluated the use of the Comaneci device for vasospasm, Tenzingplasty demonstrated superior efficacy when comparing outcomes.[7] COMMAND reported a 89.9% success rate in angiographic improvement as measured by a quartile ordinal scale and a 0.8% procedural complication rate with an occlusive thrombus resulting from treatment, whereas Tenzingplasty yielded improvement in 100% of treated segments with no procedural complications.[7] Compared to the VITAL study, Tenzingplasty was similarly superior to treatment with the NeVa device. The primary endpoint of <50% residual narrowing was achieved in 86.5% of treatments in the VITAL study, compared to 95.1% with Tenzingplasty in this study.[6] The COMMAND study reported 93.1% rate of <50% residual narrowing as reported by its ordinal scale. Mechanical dilatation with sent-retrievers or the Comaneci device may cause endothelial damage, particularly when one of these devices is dragged by pulling it through an artery while deployed or recaptured. Endothelial damage increases the risk of thromboembolic complications.[19] This was borne out with 3.2% thromboembolic complications in the VITAL study investigating the NeVa device and the 0.8% thromboembolic complication rate in the COMMAND study.[6,7] The low complication profile (0%) for Tenzingplasty is also more favorable than thromboembolic and hemorrhagic complications of angioplasty and stent-retriever-based treatment for CV reported in a recent meta-analysis.[8,20] With respect to durability, repeat endovascular mechanical treatment was required for 12.4% in the COMMAND study and 6.8% of segments in the VITAL study, whereas 3.7% of segments required repeat mechanical therapy after Tenzingplasty in the current study.[6,7] A meta-analysis of stent-retriever treatment of CV found recurrence of CV in 12.8%, although rates of retreatment with mechanical techniques comparable to those reported in COMMAND, VITAL, and the current series were not reported.[8] Whereas the VITAL study protocol only included arterial segments measuring 2-4 mm, treatment with the Tenzing devices can be performed on smaller vessel segments measuring less than 2 mm in distal branches of the MCA, ACA, or PCA, as summarized in Table 3.[6] Finally, the ability to deliver intra-arterial vasodilators through Tenzing device itself can provide super-selective infusions into difficult-to-reach distal vascular beds.

This study has several limitations, including its retrospective design, self-adjudication by operators, and short-term follow-up. This study has a modest sample size, although the number of segments treated exceeded those in the VITAL study. Additionally, heterogeneity in patient selection, treatment algorithms, and technique could introduce bias. Timing of adjuvant intra-arterial CCBs and thresholds for performing initial Tenzingplasty or determining the need for rerepeat Tenzingplasty were not standardized. This study is also limited by lack of standardized clinical outcomes reporting, which were beyond the scope of this initial technical feasibility study. Further evaluation is warranted with larger-scale, prospective investigations with harmonized inclusion criteria and treatment algorithms.

## Conclusion

Tenzingplasty appears to be a feasible, safe, and effective mechanical intervention for cerebral vasospasm, demonstrating significant improvements in caliber of treated large and medium-sized intracranial arteries with no adverse events. Tenzingplasty achieved superior efficacy with better safety compared to angioplasty with balloons, stent-retrievers, and stent-retriever-like devices. These results warrant prospective validation and may signal a potential shift in endovascular treatment for vasospasm.

## Data Availability

All data produced in the present study are available upon reasonable request to the authors

